# A Scalable Method of Applying Heat and Humidity for Decontamination of N95 Respirators During the COVID-19 Crisis

**DOI:** 10.1101/2020.04.09.20059758

**Authors:** Loïc Anderegg, Cole Meisenhelder, Chiu Oan Ngooi, Lei Liao, Wang Xiao, Steven Chu, Yi Cui, John M. Doyle

## Abstract

A lack of N95 respirators during the COVID-19 crisis has placed healthcare workers at risk. It is important for any N95 reuse strategy to determine the effects that proposed protocols would have on the physical functioning of the mask, as well as the practical aspects of implementation. Here we propose and implement a method of heating N95 respirators with moisture (85 °C, 60-85% humidity). We test both mask filtration efficiency and fit to validate this process. Our tests focus on the 3M 1860, 3M 1870, and 3M 8210 Plus N95 models. After five cycles of the heating procedure, all three respirators pass both quantitative fit testing (score of >100) and show no degradation of mask filtration efficiency. We also test the Chen Heng V9501 KN95 and HKYQ N95 finding no degradation of mask filtration efficiency, however even for unheated masks these scored <50 for every fit test. The heating method presented here is scalable from individual masks to over a thousand a day with a single industrial convection oven, making this method practical for local application inside health-care facilities.

## INTRODUCTION

While N95 filtering facepiece respirators (FFRs) were made for single use operation, the ongoing COVID-19 pandemic has caused a shortage worldwide of these masks. COVID-19, caused by the SARS-CoV-2 virus, has been shown to be very contagious and spread through aerosolized droplets. These fine droplets can remain in the air, increasing the risk of contagion to those nearby[1, 2]. This is particularly risky to healthcare workers who work with Covid patients. In order for an N95 mask to be reused more safely, the mask should be decontaminated of SARS-CoV-2 while maintaining its filtration efficiency and fit factor[3, 4]. Currently, limited information is known about SARS-CoV-2 inactivation. Initial data indicate that 70 °C dry heat for one hour was sufficient to achieve >3 log reduction in SARS-CoV2 on FFRs[5]. The addition of moisture possibly plays a role in the inactivation of some viruses[6–8]. A study found viral inactivation at 70 °C for 5 minutes[9] while in a buffered solution. SARS has also been shown to be inactivated at temperatures of 60-75 °C for 5-30 minutes in various liquid media[10–12]. Other enveloped viruses, such as H1N1 and H5N1 Influenza strains have been shown to be inactivated with moist heat at 65 °C and over 50% humidity [6–8]. These studies also found that dry heat alone was not enough to deactivate H1N1. While previous studies have looked at the effects of moist heat on N95 FFRs, these were done at substantially lower temperatures of 60-65 °C [6, 8, 13, 14]. We also note that recent CDC guidelines have indicated that moist heat is a reasonable method for reusing N95 FFRs. The resistance of *Clostridium difficile* (*C. diff*.) at elevated temperature and moisture was also studied and found temperatures of 85 °C resulted in a 5 to 6 log reduction in 15 minutes of heating in a buffer solution[15]. Here we present a scalable method of heating N95 FFRs to 85 °C, in air, with 60-85% humidity. We note that during the preparation of this manuscript, another study (with some shared co-authors) was posted on medRxiv on the filtration effects of heating under various humidity conditions and for multiple cycles of mask reuse[16].

N95 respirators are typically made of meltblown polypropylene fabric. While this naturally filters out large particles, the efficient filtration of sub-micron sized particles is due to electrostatic charge that is created in the mask fabrication process. If this charge is removed, the mask efficiency plummets, even if there are no visual signs of degradation to the mask. Thus it is critical to run filtration tests on any decontamination procedure.

There are many practical considerations which must be taken into account when trying to apply a decontamination method to hospital-scale operations. Masks should ideally be returned to the same individual for many reasons, including fit and decreased risk of spreading other diseases. Avoiding any cross contamination between masks is also important. Hospitals with limited PPE allocations would also benefit from rapid time turnaround capabilities if there is a critical shortage of N95 FFRs, ideally decontaminating a clinician’s mask between shifts.

## HEATING PROTOCOL

Here we place an N95 FFR in a 1.25 quart hard walled polypropylene container (Ziploc medium square). This ensures that both the charged layer of the mask and the container are made of the same material, which decreases the likelihood of material incompatibility. We found in our tests that the rigid container is preferable to a bag as it ensures constant volume, leading to a constant humidity and provides protection to the mask in the handling process. In each container is added a small (2.5”x2.5”) paper towel (2-ply), which is wetted with 500(*±*40) uL of water. The amount of water was initially calibrated with a fixed number of water drops. This was later replaced by a single channel pipette (Eppendorf Research Plus) capable of delivering volumes with *±*5*µ*L accuracy, which improved the repeatability of the humidity achieved (Figure 2b shows the humidity prior to this improvement.). In a hospital setting this could be replaced with an insulin syringe for example. The containers are sealed by placing the cover on and then placed in a convection oven (Despatch LAC1-38-8, 3.7 cu. Ft.). We note that a convection oven produces a stable and uniform temperature throughout the oven and heats the masks and containers quickly due to the driven warm airflow. We find that the measured oven temperature is stable to less than 1 °C regardless of the number of containers we place into this oven (1-45 containers), Figure 2a. It is important to note that a convection oven has no direct line of sight to any radiative heat sources, which would have the potential to damage the masks. For testing purposes, the lids of the containers were modified to allow a temperature and humidity sensor (SHT30 DFR0588 or SEK-SHT31-Sensors^1^) inside. We found that the humidity will drop rapidly in the presence of a small hole in the lid. The gap between the wires and the lid was thus sealed with 10 minute epoxy with a working temperature of 93 °C. For temperature verification, we also used both a K type thermocouple (readout with a SR630) and an IR thermometer (Fluke 561).

**Figure. 1.**
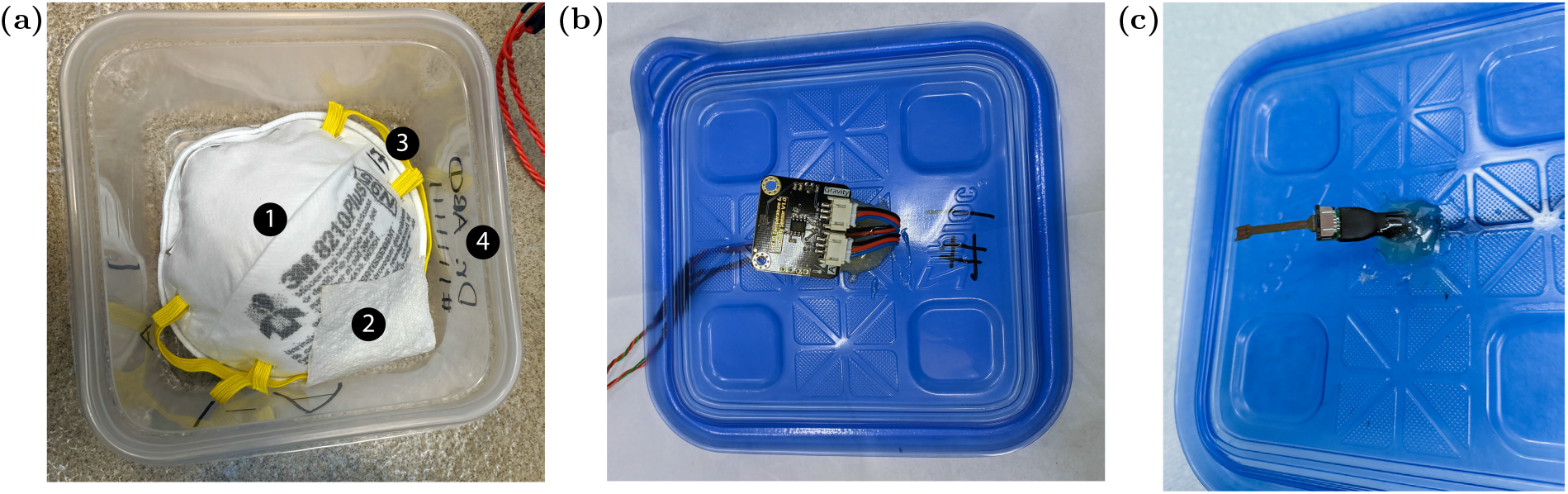
(a) Each N95 FFR (1) was placed into a plastic container with a paper towel (2) with 500 uL of water. Both the mask (3) and container (4) were labeled with a Sharpie black permanent ink marker, which would allow the mask to be identified by the correct healthcare worker and avoid cross contamination. (b/c) For testing purposes, the lid of each container was modified to have a temperature and humidity sensor ((b) SHT30 (c) SHT31).

**Figure. 2.**
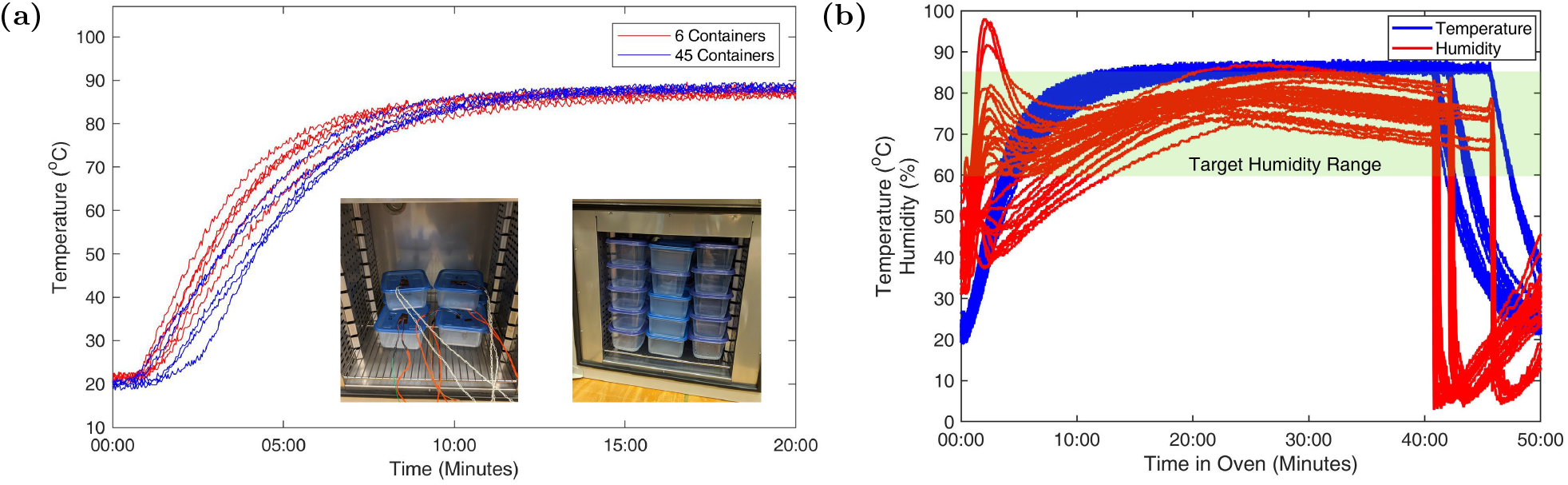
(a) Temperature rise to the set point in around 10 minutes with 45 containers vs only 6 containers, the temperature rise time is similar and final temperature are identical. Inset Left: 6 containers with humidity and temperature logging. Inset Right: 3.7 cu ft oven fully loaded with 45 containers. (b) Temperature and humidity of 3M masks which underwent filter testing during 5 heat cycles. The shaded region indicates our target range of 60-85% humidity.

Figure 3 shows the humidity as a function of time within each container for various volumes of water at various temperatures. We also find that the humidity takes between 10-15 minutes to reach the 60% level. This is an important consideration when applying this method as at least 10 additional minutes should be added to the heating cycle to achieve the desired time at target humidity. The temperature rises to within 2 °C of the target temperature in a similar time.

**Figure. 3.**
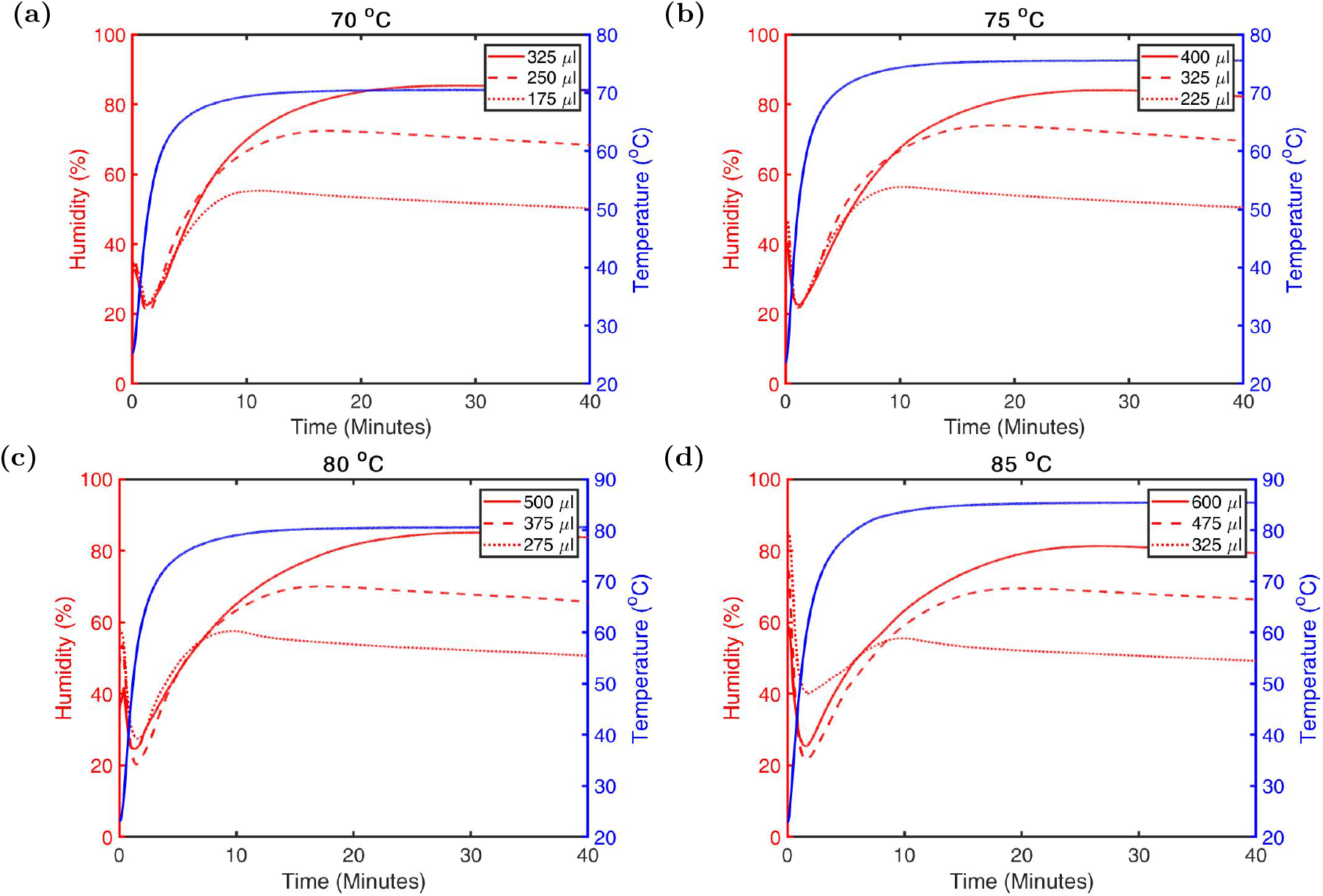
Humidity curves for various water volumes at (a) 70 °C (b) 75 °C (c) 80 °C (d) 85 °C in a 1.25 qt. container. Each humidity curve shown is an average of 3 runs.

The containers are placed into the convection oven for 40 minutes. This is due to the fact that it takes approximately 10 minutes to reach the 60% humidity level inside the container. The temperature remained stable to about 1 °C and the humidity was within a range of 60-85%. After 40 minutes, the containers were removed from the oven and for each container the lid was opened and rotated 45 degrees and placed back on top of the container, leaving air to naturally flow around the FFR. This is done while the container is still warm and we find that the processed mask is dry in under 5 minutes and the atmosphere inside the container is the same or slightly below ambient humidity after the lid is resealed. For storage in the container (or a bag, if moved out of the container) the humidity could be further reduced if desired by adding a very inexpensive small desiccant pouch into the container.

To test whether any degradation of the masks occured, we measure the filtration efficiency after five decontamination cycles. This number of cycles was chosen as previous reports, which also considered the 3M 1860, have shown that in the absence of any decontamination treatment five cycles of donning could be performed safely before fit tests began to fail consistently[17]. We tested the 3M 1860^2^ surgical N95, 3M 1870 surgical N95, 3M 8210 Plus N95, Chen Heng V9501 KN95^3^, and a HKYQ N95^4^. We perform quantitative fit test measurements after each decontamination cycle. We used a separate batch of masks for the filtration and fit tests as the masks used for fit testing must be modified to adapt to the fit testing apparatus and are worn by a user.

After the decontamination cycles the masks are inspected for any qualitative degradation. The most prominent effect observed was the delamination of the foam nose bridge on the 1870 mask, Figure 4c. Noticeable blurring of the printed labels on the 1860 and 8210 plus models, Figure 4a/b, was also seen.

**Figure. 4.**
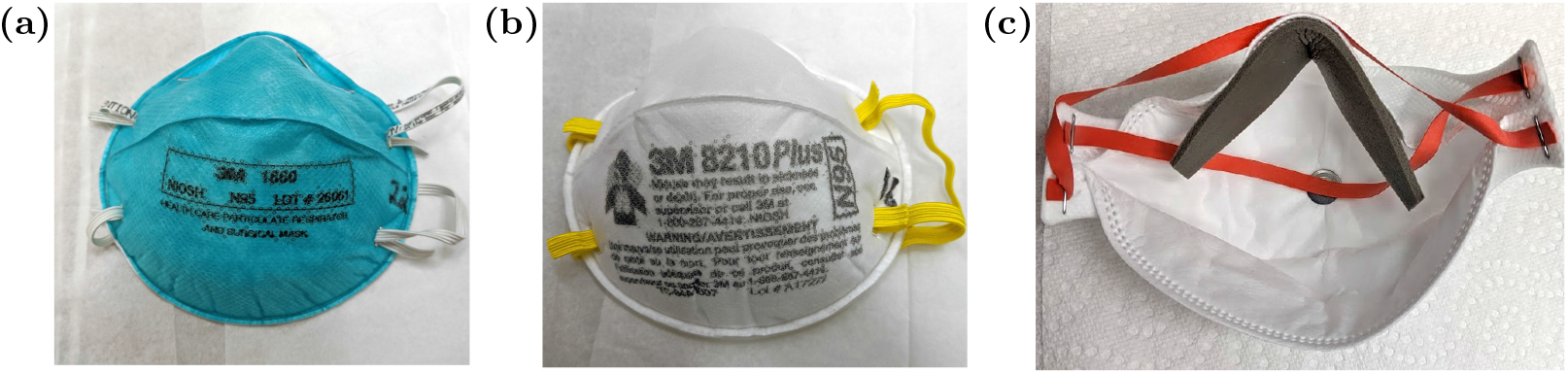
Masks after 5 heat cycles. (a) 3M 1860; Slight blurring of printed label, no other visible changes. (b) 3M 8210 Plus; Slight blurring of the printed label. (c) 3M 1870; Delamination of part of the foam nose bridge shown here after 5 cycles. The foam was reset in place prior to donning and still passed all fit tests.

## FIT TEST METHODS AND RESULTS

The fit tests were performed with a TSI PortaCount Respirator Fit Tester 8038 using NaCl particles produced by a particle generator. Each test was performed by a single user to control for variability in face structure. We used the OSHA modified ambient aerosol Condensation Nuclei Counter (CNC) Quantitative Fit Test for Filtering Facepiece Respirators. This produces a quantitative fit factor score for each of the four test components (bending over, talking, head side to side, head up and down) as well as an overall fit factor computed as the geometric mean of these four scores. A passing score is given in each category for a fit factor *≥*100, but any individual category may be failed so long as the overall fit factor is *≥*100 in order to pass the OSHA test. Additionally, the system does not provide a fit factor score greater than 200, and instead gives a maximum possible score of 200+. Each fit test sequence takes approximately two minutes and 30 seconds. For these tests each mask was punctured and fitted with a metal hose adapter to connect to the PortaCount via 3/16” ID tubing.

Prior to starting each fit test measurement, the user donned the mask and performed a qualitative user seal check to confirm that there were no detectable air leaks when forcefully exhaling. The user then made small adjustments to the fit of the mask based on the results of this seal check. After each fit test measurement the user doffed the mask to simulate the wear and tear produced through donning and doffing masks.

The protocol we performed used two masks of each type, one of which was treated as the control, and did not undergo heating, and one mask which went through five decontamination cycles. Each mask in the test group was fit tested prior to the decontamination cycles to ensure that the mask was not defective. Masks in the test group that passed this initial fit test were then run through the decontamination cycle five times. Between each cycle the test mask was removed from the container and fit tested. The control group masks were fit tested six times to correspond to the initial fit test and the fit test the test group masks would receive after each decontamination cycle. After the fifth decontamination cycle for the test group masks, we performed three additional fit tests. This was done to provide a better sampling of any variance in the user’s ability to form a tight seal with the mask and demonstrate whether there had been any degradation at the very end of the cycles.

The results of these tests are summarized in Table 1. The Chen Heng V9501 KN95 and the HKYQ N95 mask both failed the initial qualitative user seal checks, and the fit test. As a result, we declined to process these masks through the decontamination cycles and only performed further tests on the control group to determine their fit factor. The two 3M mask models, 8210 Plus and 1860, both passed the fit test on every trial, and achieved the maximum possible fit factors after five heating cycles. The 3M 1870 also passed all fit tests, however with lower fit factors after the decontamination procedure. This is most likely due to the delamination of the nose bridge foam from the mask, Figure 4c. This foam was pressed back into place by the user prior to donning.

**Table. I.** Quantitative Fit Testing results measured with a TSI PortaCount Respirator Fit Tester 8038. Red shaded cells indicate an overall fit factor <100, indicating failure of the fit test. Green shaded cells indicate successful fit tests. The cycle number indicates the number of heating cycles the test group had undergone at the time of fit testing

| Mask Type | Control or Test | Overall Fit Factor |  |  |  |  |  |  |  |
| --- | --- | --- | --- | --- | --- | --- | --- | --- | --- |
|  |  | Initial | Cycle 1 | Cycle 2 | Cycle 3 | Cycle 4 | Cycle 5 Test 1 | Cycle 5 Test 2 | Cycle 5 Test 3 |
| HKYQ N95 | Control | 28 | 9 | 6 | 9 | 19 | 26 | NA | NA |
| HKYQ N95 | Test | 5 | NA | NA | NA | NA | NA | NA | NA |
| Chen Heng V9501 KN95 | Control | 9 | 19 | 11 | 45 | 31 | 43 | NA | NA |
| Chen Heng V9501 KN95 | Test | 4 | NA | NA | NA | NA | NA | NA | NA |
| 3M 8210 Plus | Control | 127 | 200+ | 200+ | 200+ | 200+ | 200+ | NA | NA |
| 3M 8210 Plus | Test | 163 | 186 | 200+ | 200+ | 200+ | 200+ | 200+ | 200+ |
| 3M 1860 | Control | 153 | 193 | 138 | 112 | 200+ | 200+ | NA | NA |
| 3M 1860 | Test | 154 | 200+ | 200+ | 200+ | 200+ | 200+ | 200+ | 200+ |
| 3M 1870 | Control | 102 | 200+ | 200+ | 200+ | 200+ | 200+ | NA | NA |
| 3M 1870 | Test | 193 | 118 | 177 | 111 | 187 | 133 | 163 | 112 |

There is a trend in which fit tests of the 1860, and to a lesser degree the 8210 Plus and 1870, which were performed earlier in the experiment showed lower fit factors than those performed later. This is best explained by a training effect as the user gained experience with the particular mask models and identifying potential leaks during user seal checks. This effect is less clear for the other two mask models, as the user was unable to perform a satisfactory user seal check in any test.

## FILTRATION TESTING METHODS AND RESULTS

The filtration tests were performed with a TSI Inc. Automated Filter Tester 8130A using 0.26 *µ*m (mass mean diameter) NaCl as the aerosol source under a flow rate of 85 L/min. With this system we were able to determine the filtration efficiency for the challenge aerosol and the pressure drop across the mask. For these measurements we report the initial filtration efficiency as opposed to the minimum loaded efficiency, as we do not expect a significant difference in the loading curves for treated vs untreated masks.

For our filtration testing protocol we included one of each mask type in the control group and two of each type in the test group. The test group masks were each put through five heating cycles, while the control group was not heated. After the five cycles the masks were all submitted for filtration testing, the results of which are shown in Table 2.

**Table. II.** Filtration testing results. Initial filtration performance of 0.26 *µ*m (mass mean diameter) NaCl at a flow rate of 85 L/min is reported. Filtration efficiency and pressure drop shown for the test group masks is an average of the two masks of each type tested. Testing performed with a TSI Inc. Automated Filter Tester 8130A.

| Mask Type | Control Group |  | 5 Cycles at 85 °C + 60-85% Humidity |  |
| --- | --- | --- | --- | --- |
|  | Filtration Efficiency (%) | Pressure Drop (Pa) | Filtration Efficiency (%) | Pressure Drop (Pa) |
| 3M 1860 | 99.79<sup>a</sup> | 86 | 99.51 | 71.5 |
| 3M 1870 | 99.97 | 97 | 99.84 | 99.8 |
| 3M 8210 Plus | 99.78 | 84 | 99.63 | 71.5 |
| Chen Heng V9501 KN95 | 97.99 | 119.3 | 97.73 | 110.3 |
| HKYQ N95 | 98.91 | 125.7 | 98.88 | 144.3 |

<sup>a</sup> The 1860 masks used as a baseline were not expired, while, as previously noted, the masks used for the decontamination cycles were expired by 9 years.

For all four mask types studied, there was no significant drop between the control group and the test group masks. Additionally, all of the masks demonstrated a filtration efficiency above the 95% threshold for N95 respirators. The test group mask results were averaged together, but there were also no individual masks that showed an efficiency below the 95% standard. One mask of each type was also run through a 30 minute loading test to simulate the performance change when particles accumulated on the mask during the usage. During this test, the minimum efficiency for all masks remained over the 95% standard. While the pressure drop for each mask changed by up to 20%, all masks were still well below the maximum resistance to airflow specified by NIOSH ([19] 42 CFR 84.180).

## SCALABILITY

With a total cycle time of about 50 minutes, including 10 minutes for loading and unloading the oven, a 3.7 cu ft oven, as was used here, can treat approximately 1,300 masks per 24 hours under constant processing conditions. Larger convection ovens are available^5^. The equipment needed to implement this method requires the convection oven, containers, paper towels, and a pipette. This makes such a method accessible to all scales of health services.

Although the original ink labeling on the masks blurred, the labels we applied to the masks and containers with sharpie were still clear after the five cycles. This suggests a clear method for ensuring that each mask is always returned to the same user. This will be important in healthcare settings both for maintaining good fit, and because this decontamination protocol may not sterilize all potential pathogens on the masks at the temperature chosen. Additionally, the ability to label closed containers could both help to protect workers who process masks through a decontamination protocol, and to prevent cross contamination of masks during the process.

## CONCLUSIONS

This research studied the effect of five cycles of heating to 85 °C for 30 min with a relative humidity of 60-85% on a selection of N95 FFRs. We found that for all of the N95 models we investigated there was no significant difference in filtration efficacy between the test groups of masks and the untreated control masks. For the 3M 1860 and 3M 8210 Plus, there was no measurable degradation of fit after each decontamination cycle, nor relative to the control group masks. The 3M 1870 showed a slight degradation in fit factor, but still passed the OSHA quantitative fit test for every trial. As such, we found that the heating protocol described in this article is compatible with 3M models 1860, 1870, and 8210 Plus FFRs. The Chen Heng V9501 KN95 and HKYQ N95 showed no degradation in the filtration tests, but their fit factor value was in the 4-45 and 5-28 ranges respectively (where 100 is normally considered passing). Further work is still required to ensure viral inactivation of SARS-CoV-2 on FFRs under the given conditions.

## Data Availability

All relevant data is included in the manuscript text.

## ACKNOWLEDGMENTS

We would like to thank Robert Gustafson for useful discussions and the suggestion of using rigid polypropylene containers. We acknowledge and appreciate the support of the Heising-Simons foundation.

## Footnotes

1 In combination with the SEK-SensorBridge, this provides an out of the box solution with higher accuracy than the SHT30 DFR0588

2 The 1860 masks available were manufactured in 2006, with a shelf life of 5 years. Hence they were 9 years expired at the time of testing. The CDC states that model 1860 masks beyond the manufacturer shelf life have been shown to still be in accordance with NIOSH performance standards[18]. Our testing is in agreement with these findings.

3 certified to the GB2626-2006 standard

4 Zhejiang FDA Medical Device Product Certificate 20120065, certified to the GB19083 standard

5 Despatch LAC 2-18 (18 cu ft) or LBB 2-27 (27.7 cu ft) for example

